# PAD-associated genetic variants are more strongly associated with surgical intervention than premature onset

**DOI:** 10.1101/2025.05.29.25328566

**Authors:** Jiaqi Hu, Dana Alameddine, He Wang, Arya Mani, Curt Scharfe, Yong-Hui Jiang, Michael F. Murray, Cassius I. Ochoa Chaar, Andrew T. DeWan

## Abstract

Peripheral artery disease (PAD) affects over 200 million globally and often progresses without timely diagnosis, necessitating early risk identification. Recent studies have identified 19 PAD-associated single nucleotide polymorphisms (SNPs), but their associations with disease severity have not been fully explored. This study explored the association between these 19 variants and PAD severity using 6,815 PAD patients and 401,872 controls of White European ancestry from the UK Biobank. We classified PAD cases into 4 groups with increasing levels of severity based on age of PAD diagnosis and surgical status. Genetic association analysis for PAD and markers of severity was conducted using REGENIE, and a 19-variant polygenic risk score (PRS) was developed to further evaluate the associations with severity subtypes. The PRS association was then replicated using subjects from an institutional genetic program Generations (N=70 PAD cases, N=2,450 controls). From the association analysis of the 19 variants comparing four severity PAD subtypes with non-PAD controls, two SNPs (rs4722172 and rs505922) showed stronger odds ratios (ORs) with more severe subtypes. For example, the OR of rs4722172 when comparing non-premature surgical PAD with non-PAD controls was 1.08 (95% CI: 1, 1.16) while it increased to 1.18 (95% CI: 1.03, 1.36) for premature surgical PAD. Additionally, eight variants showed greater ORs for surgical compared to non-surgical PAD among premature and non-premature PAD cases respectively. Consistent with single-variant analysis, the 19-variant PRS was significantly higher in surgical groups compared to non-surgical groups (OR=1.14, 95% CI: 1.08, 1.20; p-value=5.85×10^-7^) while no significant difference was identified for non-premature and premature PAD (OR=0.98, 95% CI: 0.9, 1.07, p-value=0.64). This same pattern was observed in the subjects from Generations. Our findings demonstrate that SNPs associated with PAD development may also be associated with PAD severity as assessed by surgical intervention and age of onset (or diagnosis).

## Introduction

Peripheral artery disease (PAD) affects over 200 million people worldwide (1) and primarily impacts elderly individuals (2). It is typically diagnosed when patients present with lower extremity pain or wounds. Absence of pulses on physical exam can also alert clinicians in asymptomatic patients to obtain an ankle-brachial index test that confirms the diagnosis (3). Even though advanced stages of the disease can lead to major lower extremity amputations (1), most patients are asymptomatic and treated medically. Therefore, patients requiring surgical revascularization constitute a subgroup with more severe disease that would benefit most from early identification and risk stratification. On the other hand, patients with premature PAD who undergo revascularization at age ≤ 50 have the highest rate of amputation despite aggressive medical and surgical treatments (4). Patients with premature PAD have also been postulated to have genetic factors associated with disease onset and progression (5). Thus, the identification of PAD patients at increased risk of progression could help in developing strategies for early treatment and prevention.

Multiple lines of evidence suggest that there is a significant genetic component contributing to the development of PAD (6–8). A recent genome-wide association study (GWAS) (7) combined data from the Million Veteran Program (MVP) and the UK Biobank (UKB) identified and replicated 19 independent single nucleotide polymorphisms (SNPs) associated with the diagnosis of PAD. Additionally, the thrombophilic Factor V Leiden variant rs6025 was associated with an increased severity of PAD (7). Further, a polygenic risk score for PAD was established based on the MVP GWAS summary statistics for UKB subjects (9), and a significant association and moderate prediction accuracy were found between the PRS and the risk of PAD.

However, the association between the 19 variants and clinical indicators of advanced disease has not been fully explored. Patients with earlier onset of PAD (premature PAD) typically have a more aggressive form of the disease and are thought to be associated with genetic risk factors (4). Additionally, PAD patients that require surgical revascularization of the lower extremities may represent another group of patients with advanced disease severity and may also be associated with increased genetic risk (4).

The current study explored the association between these 19 variants and two aspects of PAD severity, premature onset and surgical lower extremity revascularization. Our hypothesis is that these 19 variants are associated with an increased PAD severity, and a polygenic risk score incorporating the 19 variants could be used to identify patients with more severe disease.

## Methods

### Study subjects

The UK Biobank (UKB) participants were enrolled between the ages of 40-69. Genotypes were assayed on either the UK BiLEVE array or the UK Biobank Axiom array with 733,332 autosomal variants overlapping between the two arrays.

For replication, we used 3,417 subjects with available genotype and phenotype data from the Yale New Haven Health System’s (YNHHS) Generations study (10). Generations is a prospective genetic program that enrolled patients in the YNHHS. Patients undergo screening for common genetic variants that predispose to diseases and make their genomic data available for research.

### Definition of clinical outcomes

In UKB, PAD cases were defined using a combination of self-report, ICD9, ICD10, and surgery codes listed in Klarin et al., 2019 (7) (Table S1). Subjects with any one of these codes were identified as PAD cases (N=6,887). PAD controls were defined as any subject not having any of the codes used to identify PAD cases (N=400,206).

PAD severity was explored using three different classification schemes.

1. PAD cases were classified into premature PAD (age of onset less than or equal to 55) and non-premature PAD (age of onset greater than or equal to 55 and less than or equal to 80). Even though prior work suggests that age 50 or younger represents premature PAD and age 60-80 is the common age of presentation, the number of patients with PAD younger than 50 was very small and the age group was expanded to 55 (4,11).
2. PAD cases were divided into surgical PAD (cases with any self-report operation or OPCS code) and non-surgical PAD (cases with no self-report operation or OPCS code).
3. PAD cases were combined from (1) and (2) above to define four mutually exclusive categories. We made two important assumptions when ranking the subtypes in terms of severity: (1) premature surgical PAD was the most severe subtype while the non-premature non-surgical PAD was the least severe subtype; and (2) the impact of age of PAD onset on severity was larger than whether receive surgery or not. These assumptions allowed us to order four subtypes with decreased severity: : a) premature surgical PAD; b) premature non-surgical PAD; c) non-premature surgical PAD; and d) non-premature non-surgical PAD.

Diabetes was defined by either ICD-10 code (UK Biobank field 41270, codes E10 and E11) or self-reported diagnosis by a doctor (field 2443 and field 20002 codes 1220, 1222 and 1223). We only consider individuals who had diabetes before the onset of PAD, and in order to maintain the sample size for PAD cases we kept those who had diabetes after PAD were coded as controls for diabetes. Diabetes was included as a covariate in later analysis.

Coronary artery disease (CAD) and cardiovascular diseases (CVD) were defined based on previous papers (12) integrating ICD 9, ICD 10, OPCS-4 and self-report records (Table S1). CAD and CVD were included in Table 1.

**Table 1.** Demographic characteristics.

| Group | N | Age at recruitment* | SEX (Male)<sup>#</sup> | BMI* | Current smoker<sup>#</sup> | Ever smoker<sup>#</sup> | Diabetes<sup>#</sup> | SBP* | DBP* | CAD<sup>#</sup> | CVD<sup>#</sup> |
| --- | --- | --- | --- | --- | --- | --- | --- | --- | --- | --- | --- |
| all | 406,991 | 56.74 (8.03) | 185,965<br>(45.7%) | 27.4<br>(4.77) | 42,830<br>(16.3%) | 144,792<br>(39.8%) | 33,276<br>(8.2%) | 139.9<br>(19.66) | 82.18<br>(10.67) | 31,014<br>(7.6%) | 209,159<br>(51.4%) |
| non-PAD | 400,206 | 56.66 (8.03) | 181,376<br>(45.3%) | 27.38<br>(4.76) | 40,978<br>(15.8%) | 141,537<br>(39.4%) | 31,685<br>(7.9%) | 139.79<br>(19.62) | 82.2<br>(10.66) | 28,361<br>(7.1%) | 203,291<br>(50.8%) |
| PAD | 6,785 | 61.53 (6.41) | 4,589<br>(67.6%) | 28.67<br>(5.31) | 1,852<br>(52.5%) | 3,255<br>(66%) | 1,591<br>(23.5%) | 145.95<br>(20.97) | 81.07<br>(11.28) | 2,653<br>(39.1%) | 5,868<br>(86.5%) |
| premature PAD | 1,047 | 53.56 (7.47) | 676<br>(64.6%) | 28.08<br>(5.56) | 330<br>(50.5%) | 393<br>(54.8%) | 105<br>(10%) | 139.53<br>(19.56) | 81.02<br>(10.81) | 335<br>(32%) | 761<br>(72.7%) |
| Non-premature PAD | 5,738 | 62.99 (4.97) | 3,913<br>(68.2%) | 28.78<br>(5.26) | 1,522<br>(52.9%) | 2,862<br>(67.9%) | 1,486<br>(25.9%) | 147.1<br>(21) | 81.08<br>(11.37) | 2,318<br>(40.4%) | 5,107<br>(89%) |
| surgical PAD | 2,534 | 61.44 (6.4) | 1,898<br>(74.9%) | 28.45<br>(4.88) | 858<br>(68.3%) | 1,278<br>(76.3%) | 573<br>(22.6%) | 147.8<br>(21.16) | 80.52<br>(11.27) | 1,021<br>(40.3%) | 2,207<br>(87.1%) |
| non-surgical PAD | 4,251 | 61.59 (6.41) | 2,691<br>(63.3%) | 28.81<br>(5.54) | 994<br>(43.7%) | 1,977<br>(60.7%) | 1018<br>(24%) | 144.86<br>(20.78) | 81.4<br>(11.28) | 1,632<br>(38.4%) | 3,661<br>(86.1%) |
| Non-premature non-surgical PAD | 3,738 | 62.88 (5.1) | 2,434<br>(65.1%) | 28.96<br>(5.49) | 873<br>(45.1%) | 1,802<br>(62.9%) | 971<br>(26%) | 146.05<br>(20.71) | 81.46<br>(11.29) | 1,499<br>(40.1%) | 3,331<br>(89.1%) |
| non-premature surgical PAD | 2,000 | 63.19 (4.7) | 1,479<br>(74%) | 28.43<br>(4.77) | 649<br>(69%) | 1,060<br>(78.5%) | 515<br>(25.8%) | 149.07<br>(21.42) | 80.37<br>(11.47) | 819<br>(41%) | 1,776<br>(88.8%) |
| premature non-surgical PAD | 513 | 52.21 (7.16) | 257<br>(50.1%) | 27.67<br>(5.77) | 121<br>(35.8%) | 175<br>(44.6%) | 47<br>(9.2%) | 136.01<br>(19.11) | 80.98<br>(11.17) | 133<br>(25.9%) | 330<br>(64.3%) |
| premature surgical PAD | 534 | 54.86 (7.53) | 419<br>(78.5%) | 28.51<br>(5.32) | 209<br>(66.1%) | 218<br>(67.1%) | 58 (10.9%) | 142.94<br>(19.41) | 81.06<br>(10.45) | 202<br>(37.8%) | 431<br>(80.7%) |

PAD: peripheral artery disease; SBP: systolic blood pressure; DBP: diastolic blood pressure; CAD: coronary artery disease; CVD: cardiovascular disease.
\*: mean (standard deviation); <sup>#</sup>: count (proportion%).

#### Generations clinical definitions

Similarly, patients in the Generations study were defined as having PAD based on ICD 10 codes and current procedural terminology (CPT) codes related to lower extremity revascularization. (Table S2)

### Quality control

#### UKB

UKB phenotype-independent quality control was performed as previously described (13). Subjects were selected based on those who self-defined and were genetically confirmed as White British (field 22006). This dataset was expanded to include all White Europeans by using self-reported ancestry and pre-computed PCs (field 22009). First, the mean and covariance were calculated for 40 PCs from the genetically confirmed White British subjects (N=409,615). For each subject, the Mahalanobis distance from the empirical PC distribution was calculated. Using a Mahalanobis distance threshold that captures 94% (N=459,438) of subjects, we captured all genetically confirmed White British subjects (N=409,615) in addition to 49,823 non-British White subjects. These subjects were then subject to additional phenotype-independent QC including call rate ≤99% (N=46,394), no phenotype data available (N=95), genetic and reported sex mismatch (N=417), sex chromosomes non-XX XY (N=512), outliers in heterozygosity/missing rate (N=1,095), heterogeneity rate > mean±3*standard deviations (N=2,064), and withdrawn from the study (N=20) (see Figure S1). Following the construction of the PAD phenotype and covariates, we excluded additional subjects, including, subjects missing PAD status (N=31), PAD cases with age of diagnosis larger than 80 (N=102), and subjects with missing covariates (N=1,717) (see Figure S1). We included 406,991 subjects from UKB in our analysis.

Variants were initially quality controlled in a phenotype-independent manner as described previously (13). Variants were excluded if they had a call rate ≤ 99%, Hardy-Weinberg equilibrium (HWE) p-value < 5×10^-8^ and minor allele frequency (MAF) ≤ 0.001 (N=47,489). We did further QC on our case-control data in a phenotype-dependent manner. For directly genotyped variants, we excluded those with 1) call rate ≤99% in cases or controls respectively (N=1,815); and 2) HWE p-value < 5×10^-8^ in controls (N=2,381) (Figure S2).

For imputed variants, those with 1) INFO Score < 0.8; and 2) MAF < 0.001 were excluded. The numbers of analyzed SNPs were 13,417,818 for non-premature non-surgical PAD, 13,416,573 for non-premature surgical PAD, 13,416,613 for premature non-surgical PAD, and 13,416,913 for premature surgical PAD respectively.

#### Principal components analysis (PCA)

Using the set of directly genotyped variants, the variants were LD pruned using the default parameters in a 50 variant window so that none have a pairwise r^2^ of greater than 0.2 and shifting by 5 variants after each step in Plink-1.90 (14) which resulted in nearly 237,200 variants. PCA was performed on these variants using the smartPCA program in EIGENSOFT-7.2.1(15).

#### Generations

After initial quality control removing 9 subjects with a call rate < 95%, we inferred the genetic ancestry for the remaining 3,408 subjects by comparing them to the 503 confirmed White European subjects from 1000 Genome Phase 3 (16) using a principal component analysis. We then calculated the Mahalanobis distance across the top 10 PCs. Generations subjects who clustered within the 1000 Genome White Europeans were retained in our replication sample (N=2,520).

### Statistical analysis

#### Association analysis (GWAS)

Despite our interest in a small subset of variants, we performed the association analysis on the full set of variants to account for any relatedness and population stratification in the dataset.

Univariate association analysis was performed on the full set of imputed and directly genotyped variants using linear and logistic regression models implemented in REGENIE (17). REGENIE implements a multi-stage procedure that starts by fitting a null model with whole genome regression, approximates to the linear mixed model, using ridge regression on the directly genotyped variants (N=524,010) to estimate the polygenic effects parameter to account for relatedness and population stratification. Then, it uses the full set of imputed variants to estimate the genetic association of the phenotype using a leave-one-chromosome-out procedure. For binary traits, Firth regression is used to ensure that the results are well-calibrated in the presence of low-frequency variants and unbalanced case-control ratios, both of which are present in this analysis. REGENIE implements an approximate Firth regression so that this approach can be run on a genome-wide scale. To adjust for confounding and potential bias due to population stratification, sex, age at recruitment, and top 10 PCs were included as covariates.

#### PRS development

We integrated the 19 previously identified variants into a polygenic risk score (PRS) to analyze their aggregate effects on PAD severity. The PRS was calculated as the weighted sum of the number of minor alleles for each subject. The weights, in other words, the beta coefficients, for each variant were extracted from the Million Veteran Project (MVP) European GWAS summary statistics of PAD, calculated using 24,009 PAD cases and 150,984 non-PAD controls (7) and obtained from the database of Genotype and Phenotype (dbGaP; accession number 32518). The final PRS was standardized to mean zero and a standard deviation of one among study subjects.

#### PRS analysis

The association between the PRS and PAD, as well as the severity subgroups, was assessed through logistic regression models. The covariates included age at recruitment, sex, top 10 PCs, smoking status (never, former, and current), and diabetes. To further evaluate the prediction performance, we used a five-fold cross-validation approach to calculate the area under the curve (AUC). Specifically, the data was randomly divided into training and testing sets with a ratio of 4:1. Logistic regression models were fitted in the training set and the risk for subjects in the testing set was estimated through the same model. The AUC was computed to measure the prediction accuracy. The process was repeated five times and the average AUCs were computed. The significance level was set to 6.58×10^-4^ (0.05/(19 variants *4 comparison groups)) for the single variant analyses and 1.79×10^-3^ (0.05/(14 comparison groups *2 models)) for PRS analyses. Odds ratios (ORs) were reported per standard deviation and the adjusted ORs were calculated using model incorporating covariates. In addition to REGENIE and PLINK, some quality control and statistical analysis as well as visualization of association results were done by R 4.2.

#### Sensitivity analysis

We developed a genome-wide PRS via SDPR (18) as a sensitivity analysis. Specifically, we used 927,285 SNPs present in MVP GWAS summary statistics, UKB imputed genotype data after quality control, and the LD reference data provided by SDPR (1 million HapMap3 SNPs estimated from 503 1000 Genome European samples). The weights were inferred through SDPR and the SRPR PRS was further calculated for UKB subjects included for 19-variant PRS analysis via PLINK 1.90. The same analyses including associations with PAD severity subtypes and prediction accuracy were conducated using the SDPR PRS.

### Ethical approval (UKB)

This research was conducted using the UK Biobank Resource (application number 32285) (19). The UK Biobank study was conducted under generic approval from the National Health Services’ National Research Ethics Service. The present analyses were conducted in accordance with the Declaration of Helsinki and approved by the Human Investigations Committee at Yale University (2000026836 for UK Biobank and 2000024015 for Generations). The patients/participants provided their written informed consent to participate in this study.

### Results Demographics

In total, 406,991 participants were eligible for the study, of which 6,785 were patients with PAD (cases), and 400,206 were subjects that did not have PAD (controls). PAD cases were then subset into the four different severity subgroups based on age of diagnosis and surgical revascularization. The characteristics of the cases and controls, as well as case subgroups, are presented in Table 1. Compared to control subjects with no PAD, patients with PAD were more likely to be male (67.63% vs. 45.32%, p-value<2×10^-16^), older at recruitment (61.53 vs. 56.66, p-value<2×10^-16^), have higher systolic blood pressure (SBP) (145.95 vs. 139.79, p-value<2×10^-16^) and body mass index (BMI) (28.67 vs. 27.38, p-value<2×10^-16^), and have a higher proportion of current (52.46% vs. 15.84%, p-value<2×10^-16^) and former smokers (65.98% vs. 39.4%, p-value<2×10^-16^). Higher proportion of patients with diabetes could be found in PAD cases than in controls (23.45% vs. 7.92%, p-value<2×10^-16^). We focused on the previously identified 19 variants associated with PAD(7). Comparing PAD cases to non-PAD controls, we found similar effect estimates and at least nominal significance for 15 of the 19 variants (Table 2).

**Table 2.**
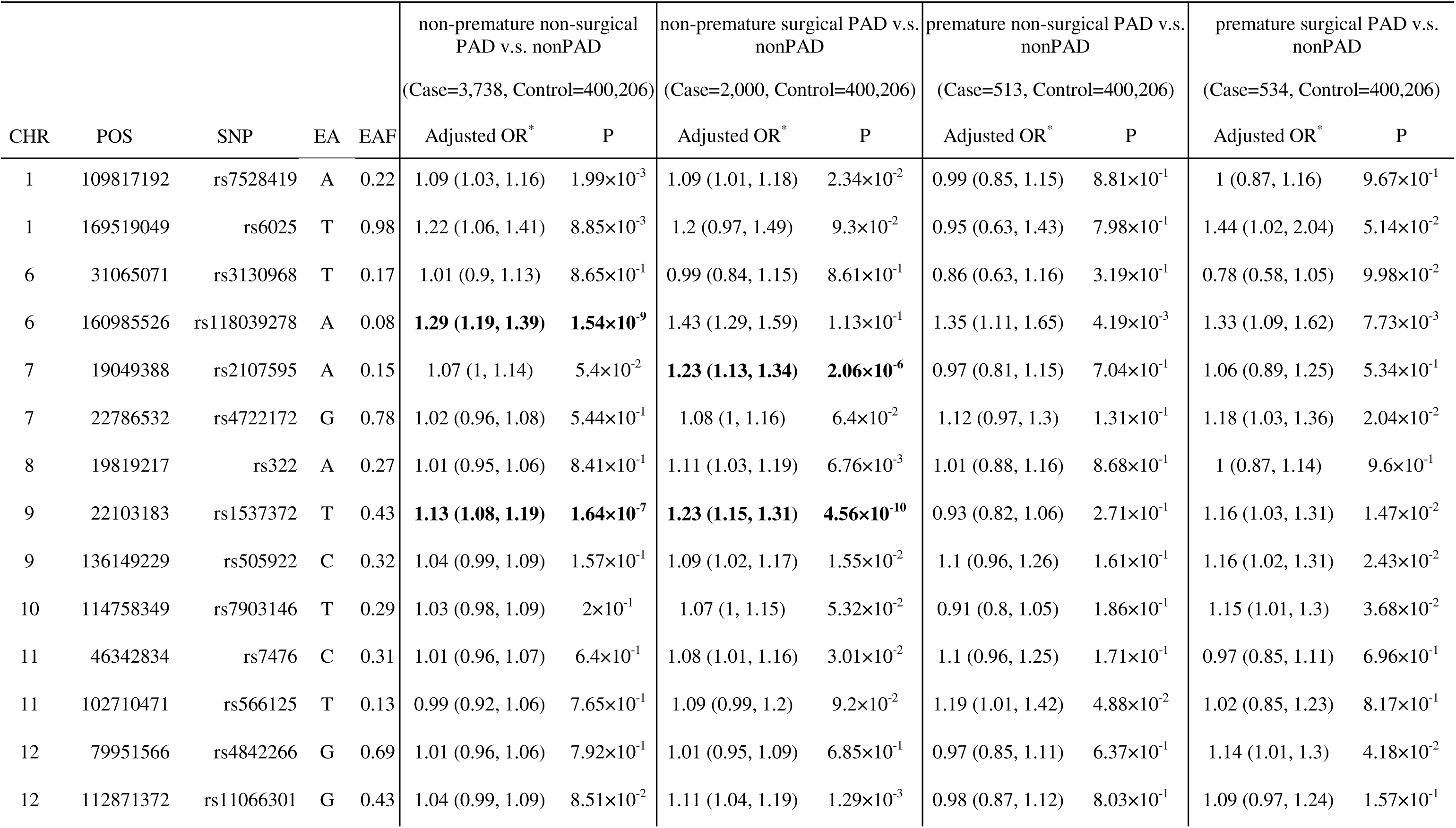
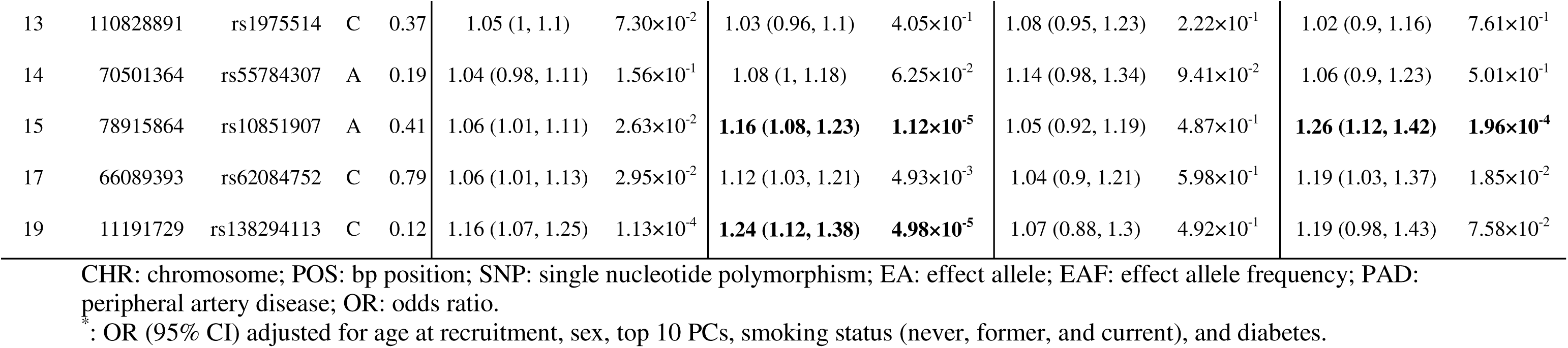
Associations between 19 variants and four severity PAD subgroups compared with non-PAD controls.

### Single-variant association analysis

We then explored the hypothesis that some or all of these 19 variants are associated with PAD severity by testing the association between each of the four PAD subgroups compared to the control subjects with no diagnosis of PAD. We hypothesized that early age of onset (≤55) and surgical intervention for PAD were indicators of increased severity and thus classified the four PAD subgroups from least to most severe as: 1) non-premature, non-surgical PAD (N=3,738); 2) non-premature, surgical PAD (N=2,000); 3) premature, non-surgical PAD (N=513); and 4) premature, surgical PAD (N=534).

Two variants (rs4722172 and rs505922) showed increased adjusted ORs with increasing severity (Figure 1 and Table 2). rs4722172 had adjusted ORs of 1.02 (95% CI: 0.96, 1.08; p-value=5.44×10^-1^) for non-premature, non-surgical PAD; 1.08 (95% CI: 1, 1.16; p-value=6.4×10^-2^) for non-premature, surgical PAD; 1.12 (95% CI: 0.97, 1.3; p-value=1.31×10^-1^) for premature, non-surgical PAD; and 1.18 (95% CI: 1.03, 1.36; p-value=2.04×10^-2^) for premature, surgical PAD. rs505922 had adjusted ORs of 1.04 (95% CI: 0.99, 1.09; p-value=1.57×10^-1^) for non-premature, non-surgical PAD; 1.09 (95% CI: 1.02, 1.17; p-value=1.55×10^-2^) for non-premature, surgical PAD; 1.10 (95% CI: 0.96, 1.26; p-value=1.61×10^-1^) for premature, non-surgical PAD; and 1.16 (95% CI: 1.02, 1.31; p-value=2.42×10^-2^) for premature, surgical PAD. Another 8 variants showed larger ORs for surgical compared to non-surgical PAD stratified by age of onset (Figure S3). For example, rs1537372 was associated with an increased risk for surgical PAD (adjusted OR=1.16, 95% CI: 1.03, 1.31; p-value=1.47×10^-2^ [premature] and adjusted OR=1.23, 95% CI: 1.15, 1.31; p-value=4.56×10^-10^ [non-premature]) while the adjusted ORs for non-surgical PAD were lower (adjusted OR=0.93, 95% CI: 0.82, 1.06; p-value=2.7×10^-1^ [premature] and 1.13, 95% CI: 1.08, 1.19; p-value=1.64×10^-7^ [non-premature]). Of note, after Bonferroni correction (p-value < 6.58×10^-4^), three SNPs were significantly associated with an increased risk for non-premature non-surgical PAD (rs118039278, rs1537372, and rs138294113), and they were also significantly associated with non-premature surgical PAD along with other two variants (rs2107595 and rs10851907). rs10851907 was significantly associated with premature surgical PAD as well.

**Figure 1.**
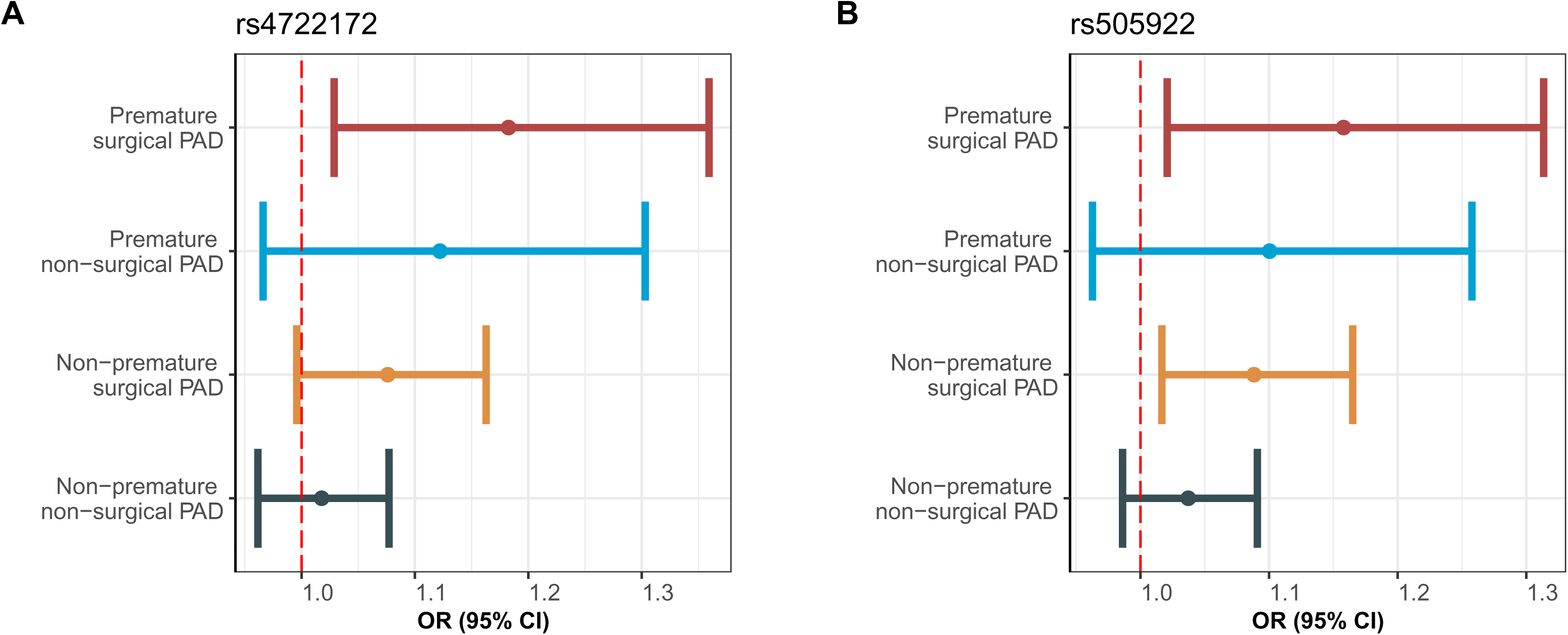
Association analysis for rs4722172 and rs505922 comparing four severity groups and non-PAD controls. Association analysis of 19 variants in four severity PAD subgroups compared with non-PAD controls revealed that two variants, rs4722172 (A) and rs505922 (B), showed increasing odds ratios (ORs) across four subgroups.

### PRS analysis

To investigate the aggregate effects of these 19 variants, we calculated a PRS for 406,991 subjects. Using the Bonferroni correction threshold (p-value<1.79×10^-3^), The standardized PRS was higher in PAD cases than in non-PAD controls (Figure S4) and significantly associated with PAD status (OR=1.23, 95% CI: 1.2, 1.26; p-value=1.91×10^-67^) and the significance remained after adjusting for covariates (Figure 2). Additionally, the PRS was significantly associated with all four severity PAD subtypes compared to non-PAD controls (Figure 2). The adjusted ORs were 1.17 (95% CI: 1.14, 1.21; p-value=1.32×10^-22^), 1.34 (95% CI: 1.29, 1.4; p-value=3.93×10^-41^), 1.12 (95% CI: 1.02, 1.22; p-value=1.3×10^-2^), 1.32 (95% CI: 1.21, 1.43; p-value=6.7×10^-11^) for non-premature non-surgical PAD, non-premature surgical PAD, premature non-surgical PAD, and premature surgical PAD, respectively. The adjusted ORs for surgical PAD were higher than adjusted ORs for non-surgical PAD stratified by age of onset while the adjusted ORs for premature PAD were lower than adjusted ORs for non-premature PAD stratified by surgery. We further combined subgroups into surgical PAD (n=2,534) vs. non-surgical PAD (N=4,251) and premature PAD (N=1,047) vs. non-premature PAD (N=5,738) and analyzed the associations between PRS and specific combined severity groups. PRS was significantly associated with higher risk for all four combined severity subtypes (Table 3). The largest adjusted OR could be found for surgical PAD (adjusted OR=1.34, 95% CI: 1.29, 1.39; p-value=5.45×10^-50^) and it was higher than the adjusted OR for non-surgical PAD (adjusted OR=1.17, 95% CI: 1.13, 1.20; p-value=1.58×10^-23^) and did not have overlapping 95% CIs. Further test showed a significantly higher PRS for surgical PAD than the non-surgical PAD (adjusted OR=1.14, 95% CI: 1.08, 1.20; p-value=5.85×10^-7^). The adjusted ORs for premature PAD (adjusted OR=1.21, 95% CI: 1.14, 1.29; p-value=1.84×10^-10^) and non-premature PAD (adjusted OR=1.23, 95% CI: 1.2, 1.27; p-value=2.63×10^-55^) were similar with overlapping 95% CIs. No significant association between PRS and premature PAD compared with non-premature PAD was identified (adjusted OR=0.98, 95% CI: 0.9, 1.07, p-value=0.64).

**Figure 2.**
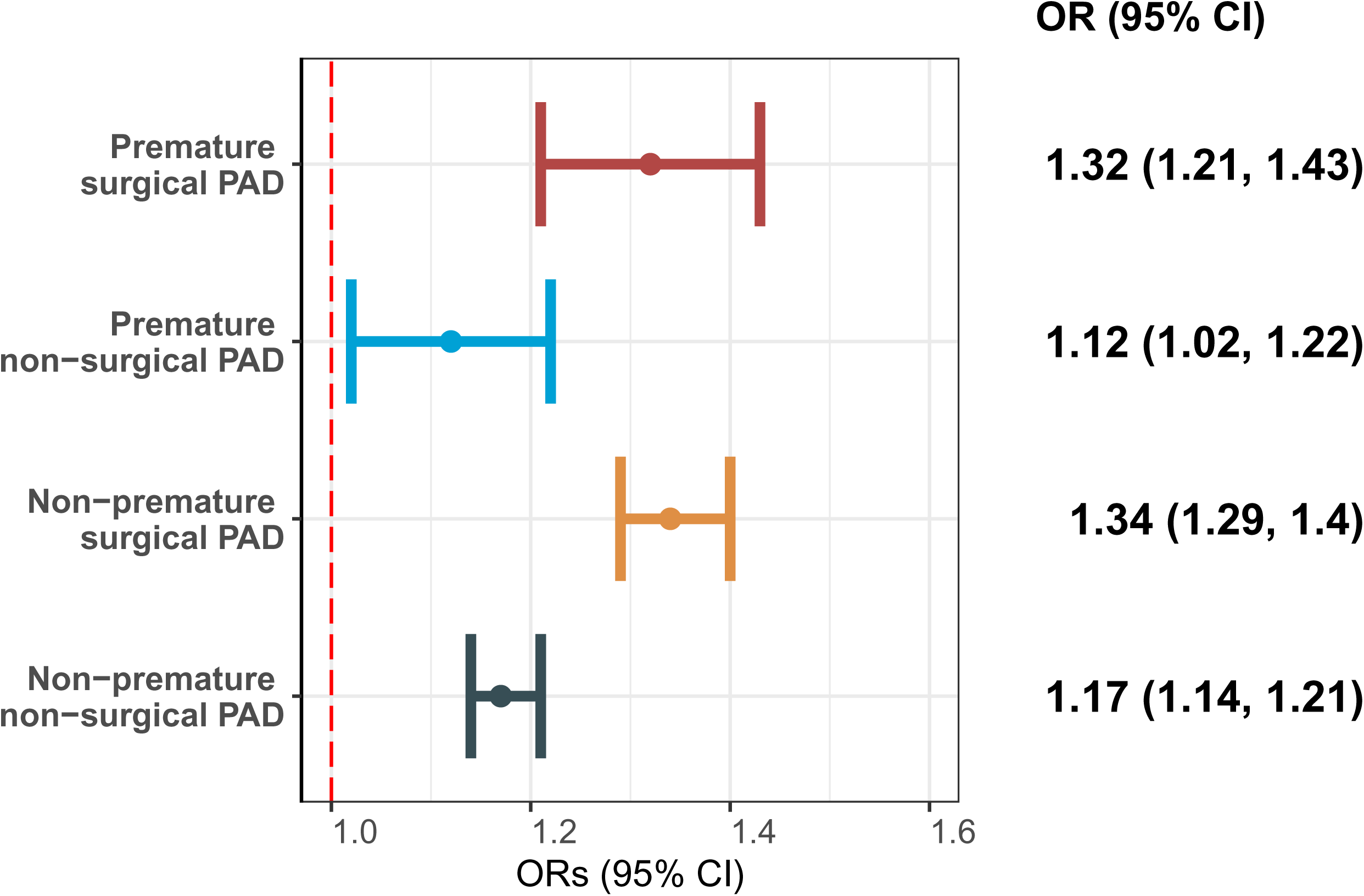
Association between PRS and four severity PAD subgroups compared with non-PAD controls. Using effect sizes from MVP summary statistics, we developed a 19-variant PRS and evaluated its associations with PAD severity adjusting for covariates. The PRS was significantly associated with an elevated risk for all four severity subgroups compared to non-PAD controls, with the highest OR observed in the premature surgical PAD subgroup and the second in the non-premature surgical PAD subgroup. These ORs were significantly higher compared to the respective non-surgical counterparts. However, no significant differences were found between premature and non-premature PAD groups for ORs in both surgical and non-surgical groups.

**Table 3.** PRS associations with surgical status and age of onset compared with non-PAD controls.

| Phenotypes | OR (95% CI) | P-value |
| --- | --- | --- |
| Surgical PAD | 1.34 (1.29, 1.39) | $5.45 \times 10^{-50}$ |
| Non-surgical PAD | 1.17 (1.13, 1.2) | $1.58 \times 10^{-23}$ |
| Premature PAD | 1.21 (1.14, 1.29) | $1.84 \times 10^{-10}$ |
| Non-premature PAD | 1.23 (1.2, 1.27) | $2.63 \times 10^{-55}$ |

539 PRS: polygenic risk score; PAD: peripheral artery disease; OR: odds ratio.

Additionally, we assessed whether the PRS could accurately predict the risk for PAD and the severity subtypes by assessing the area under curve (AUC). The PRS showed a low AUC (0.56 95% CI: 0.54, 0.57) for PAD. As for severity subtypes, the highest AUC could be found for surgical PAD (0.59 95% CI: 0.56, 0.62 [non-premature] and 0.58 95% CI: 0.52, 0.63 [premature]) (Figure S5).

### PRS replication analysis

We repeated the PRS analysis among Generations subjects to replicate our findings of the association between increased PRS and risk for surgical and non-surgical PAD. A total of 14 non-surgical PAD, 56 surgical PAD, and 2,446 non-PAD controls were analyzed. Based on the Bonferroni correction threshold (p-value<1.79×10^-3^), the PRS was significantly associated with a higher risk of surgical PAD (adjusted OR=1.54, 95% CI: 1.18, 2.01, p-value=1.29×10^-3^) while the association between the PRS and non-surgical PAD was not significant (adjusted OR=0.83, 95% CI: 0.47, 1.46, p-value=0.52). No significant association was found for PRS and surgical PAD compared with non-surgical PAD (adjusted OR=7.32, 95% CI: 0.85, 62.67; p-value=6.92×10^-2^). To sum up, we successfully replicated the associations between PRS and a higher risk of surgical PAD compared to non-PAD controls but failed to replicate the associations between PRS and non-surgical PAD compared with non-PAD controls and the higher PRS found in surgical than in non-surgical PAD.

#### PRS sensitivity analysis

The genome-wide PRS was developed for UKB European subjects through SDPR. We conducted association analyses with PAD severity for the SDPR PRS, and the SDPR PRS was significantly higher in surgical than in non-surgical PAD cases (adjusted OR=1.10, 95% CI: 1.04, 1.16; p-value=2.91×10^-4^). Futhermore, we evaluated the prediction accuracy of the SDPR PRS via AUC. The highest AUC was identified when comparing non-premature surgical PAD with non-PAD controls (0.61, 95% CI: 0.58, 0.64), and this observation was similar to the 19-variant PRS. Comparing the AUCs in each severity comparison group with the 19-variant PRS, we found the AUCs estimated using the SDPR PRS were slightly higher than the 19-variant PRS but the 95% CIs were overlapped, suggesting no significant differences (Table S3). These results indicated that our 19-variant PRS can capture most of the genetic risk that was estimated using a genome-wide PRS.

## Discussion

This study highlights the association of genetic variants with the severity of PAD especially in patients requiring surgical revascularization. Under the assumption of increasing severity across these subgroups, we observed that 2 of the 19 variants showed increased adjusted ORs.

Subsequently, a 19-variant PRS was significantly higher in surgical subjects compared to non-surgical cases when stratified by age of onset. However, there was no significant difference in PRS between non-premature and premature PAD cases. The significant association between PRS and surgical PAD was replicated in an independent dataset.

In this study, we developed a PRS incorporating 19 variants identified by Klarin et al. (2019) with effect sizes from the MVP GWAS summary statistics. This PRS demonstrated a significant association with the risk of PAD (adjusted OR 1.23, 95% CI: 1.2, 1.26). This finding aligns with a prior study that reported an adjusted OR of 1.44 (95% CI 1.41, 1,46) (9). The difference in the magnitude of estimated effects could be attributed to several factors: (1) our analysis was restricted to individuals with genetically confirmed European ancestry, whereas the previous research encompassed individuals from varied genetic backgrounds, including African descent. Despite adjusting for the top five PCs, the previous study may suffer from residual confounding due to population stratification, potentially biasing the estimated adjusted OR; (2) Our model incorporated two additional covariates, smoking status and diabetes, which were not considered in the prior study. This inclusion could influence the estimated association; (3) The previous study employed a more sophisticated method for inferring PRS weights and included more SNPs. While this approach enhanced power and captured more signals, it may also have introduced additional noise into the PRS estimation; (4) The 19 variants were replicated using UKB data and constructing PRS with these variants in UKB might suffer from sample overlapping issue, which might bias the association estimates.

A significant association was identified and replicated between the 19-variant PRS and surgical intervention. Even though the majority of patients with diagnosis of PAD are asymptomatic, it is estimated that 10% will progress and require lower extremity revascularization. Thus, the surgical group in the current analysis represents patients with more severe form of PAD, that was correlated with the PRS indicating a link between the 19 variants and the severity of PAD. This observation also highlights the potential and non-immediate utility of aggregating genetic variants to predict the severity of PAD to provide insights for early prevention and possibly more aggressive medical therapy upfront. However, despite its association with surgical intervention, the PRS did not show a significant difference between premature and non-premature PAD cases, which might be explained by two factors. First, the PRS was developed from 19 variants, of which only 2 SNPs exhibited higher adjusted ORs in premature PAD cases than in non-premature cases (Figure 1). Therefore, these 19 variants might not sufficiently capture the genetic distinctions between premature and non-premature PAD. Second, the relatively small sample sizes of premature PAD cases could have limited the statistical power of our analyses, thus affecting our ability to detect significant differences.

This study has several limitations. First, we made two important assumptions when ranking the four severity subtypes, and further evidence was needed to verify the assumptions. Second, the investigation for associations between 19 SNPs and PAD severity subtypes was designed and conducted as a hypothesis-generating analysis. Consequently, further research is necessary to test the hypothesis that rs4722172 and rs505922 are associated with an increased trend across PAD severity subgroups. Third, some patients with advanced PAD may require surgery but may not be suitable surgical candidates and may have been grouped into the non-surgery PAD groups.

However, with the advances of minimally invasive endovascular surgery the proportion of symptomatic patients who are not offered revascularization is likely very small. At last, the sample sizes for subgroups, particularly the premature surgical PAD group, were limited in both the discovery and replication datasets. Future studies with larger sample sizes should be conducted to increase the power to dissect these associations with phenotypic subgroups of PAD cases. Finally, our study focused on individuals with European ancestry. It is crucial for future research to extend these analyses to other ancestry groups.

## Conclusion

This study demonstrated that the PRS was associated with an increased risk of surgical revascularization in patients with PAD. Future research efforts with larger sample sizes should be conducted to identify variants associated with premature PAD and incorporated into a PRS.

## Data Availability

Summary statistics for UKB generated during and/or analyzed during the current study are available in Table 2. Summary statistics for MVP are available by a request directly to dbGaP. The summary statistics from Generations for the 19 variants are available upon request.

## Consent of publication

All authors agree to publish.

## Authors’ Contributions

JH, DA, CIOC, and AD contributed to the study conception and design. Data requests and analysis of the discovery datasets were performed by JH, DA, CIOC, and AD. HW, AM, CS, YHJ, and MFM contributed to the data collection and replication of the replication dataset. The first draft of the manuscript was written by JH and AD, and all authors commented on the manuscript. All authors read and approved the final manuscript.

### Acknowledgment

The authors would like to thank the researchers and participants of the United Kingdom Biobank. All data was accessed as part of project 32285 from the United Kingdom Biobank. We thank the Yale Center for Research Computing for the use of the McCleary High Performance Computing cluster.

## Clinical trial number

Not applicable.

