## Supplementary material for "PAD-associated genetic variants are more strongly associated with surgical intervention than premature onset": Table S1-3, Figure S1-5

### Table S1 Definitions for PAD, diabetes, CAD, and CVD in UKB

|  | Self-report disease | ICD9 | ICD10 | Self-report operation | OPCS-4 | Others |
| --- | --- | --- | --- | --- | --- | --- |
| PAD | 1067, 1087, 1088 | 4400, 4402, 4438, 4439 | I70.00, I70.01,  I70.20, I70.21,  I70.80, I70.90, I73.8, I73.9 | 1102, 1108, 1440 | X09.3, X09.4, X09.5 L21.6, L51.3, L51.6, L51.8, L52.1, L52.2, L54.1, L54.4, L54.8 L59.1, L59.2, L59.3, L59.4, L59.5, L59.6, L59.7, L59.8, L60.1, L60.2, L63.1, L63.5, L63.9, L66.7 |  |
| Diabetes | 1220, 1222, 1223 |  | E10, E11 |  |  | 2443 |
| CAD | 1075 | 4109, 4119, 4129, 429.79 | I21-I23, I24.1, I25.2 | 1075, 1095 | K40, K41, K45, K49, K50.2, K75 |  |
| CVD | 1074, 1075, 1082, 1583 | 4109, 4119, 4129, 4139, 4140, 4141, 4148, 4149, 4349, 4369 | G45, I20-I25, I63, I64 | 1070, 1071, 1095, 1105, 1109, 1514 | K40-46, K47.1, K49, K50, K75 |  |

PAD: peripheral artery disease; CAD: coronary artery disease; CVD: cardiovascular disease; UKB: UK Biobank; ICD9/10: International Classification of Diseases, Ninth/Tenth Revision.

### Table S2 Definitions for PAD in GENERATION

|  | ICD9 | ICD10 | CPT |
| --- | --- | --- | --- |
| PAD | 249.7, 249.71, 250.7, 250.71, 250.72, 250.73, 440.2, 440.20, 440.21, 440.22, 440.23, 440.24, 440.29, 440.4, 443.9 | I70.211, I70.212, I70.213, I70.221, I70.222, I70.223, I70.231, I70.232, I70.233, I70.241, I70.242, I70.243, I70.261, I70.262, I70.263, I70.92, I73.9 | 34832, 35302, 35303, 35304, 35305, 35306, 35331, 35351, 35355, 35361, 35363, 35371, 35372, 35452, 35454, 35456, 35459, 35470, 35472, 35473, 35474, 35481, 35482, 35483, 35485, 35491, 35492, 35493, 35495, 35521, 35533, 35537, 35538, 35539, 35540, 35548, 35549, 35551, 35556, 35558, 35563, 35565, 35566, 35570, 35571, 35583, 35585, 35587, 35621, 35623, 35637, 35638, 35646, 35647, 35651, 35654, 35656, 35661, 35663, 35665, 35666, 35671, 37220, 37221, 37222, 37223, 37224, 37225, 37226, 37227, 37228, 37229, 37230, 37231, 37232, 37233, 37234, 37235 |

PAD: peripheral artery disease; ICD9/10: International Classification of Diseases, Ninth/Tenth Revision; CPT: current procedural terminology.

**Table S3 AUC for 19-variant and SDPR PRS**

| Outcome | AUC estimated by 19-variant PRS (95% CI) | AUC estimated by SDPR PRS (95% CI) |
| --- | --- | --- |
| PAD | 0.56 (0.54, 0.57) | 0.59 (0.57, 0.6) |
| Non-premature non-surgical PAD | 0.54 (0.52, 0.57) | 0.57 (0.55, 0.59) |
| Non-premature surgical PAD | 0.59 (0.56, 0.62) | 0.61 (0.58, 0.64) |
| Premature non-surgical PAD | 0.53 (0.48, 0.59) | 0.55 (0.49, 0.61) |
| Premature surgical PAD | 0.58 (0.52, 0.63) | 0.6 (0.55, 0.66) |

AUC: area under the ROC curve; PRS: polygenic risk score; PAD: peripheral artery disease.

**Figure S1 Flowchart of quality control for individuals**

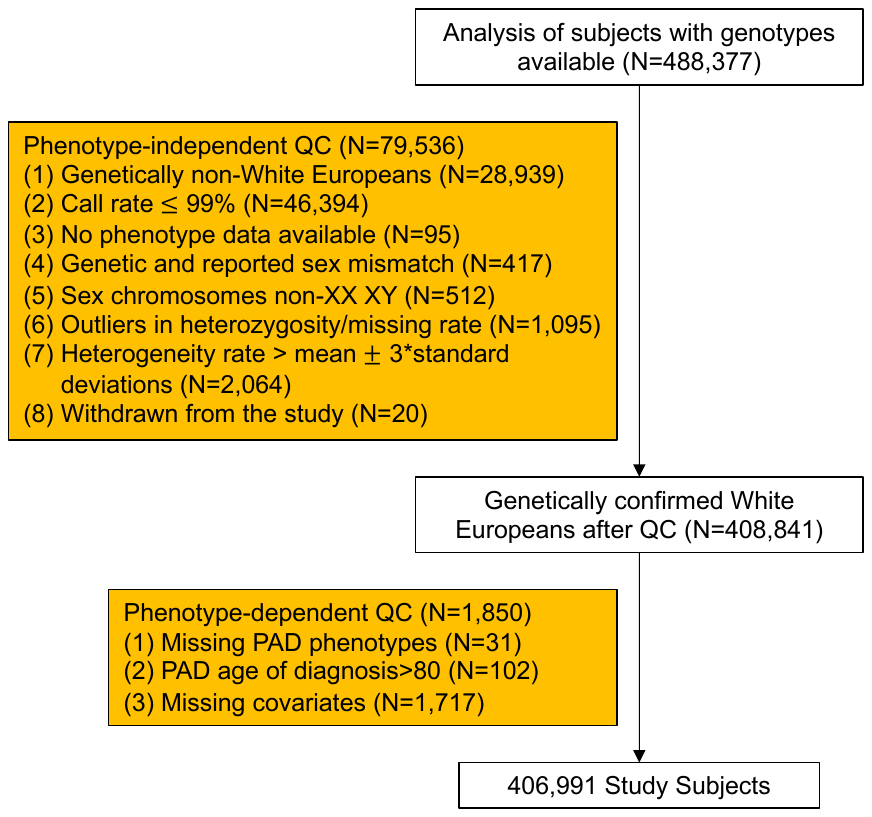

The flowchart of quality control on UKB subjects. Individuals were removed due to criteria in the yellow boxes and the final sample size for study was 406,991.

**Figure S2 Flowchart of quality control for SNPs**

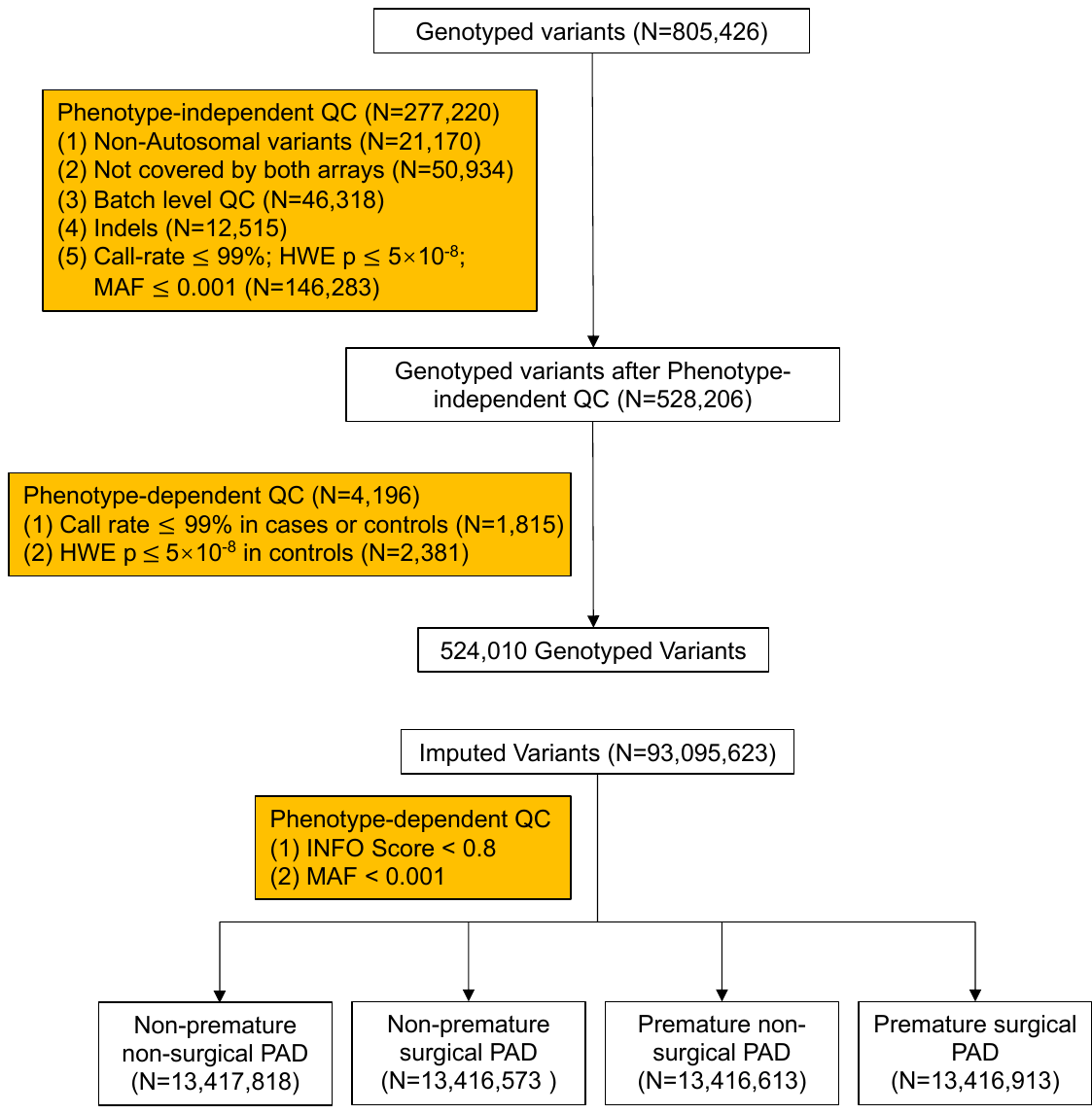

The flowchart of quality control on UKB raw and imputed genotype. SNPs were removed due to the criteria in yellow boxes.

**Figure S3 Associations for seventeen variants between four severity groups compared with non-PAD controls.**

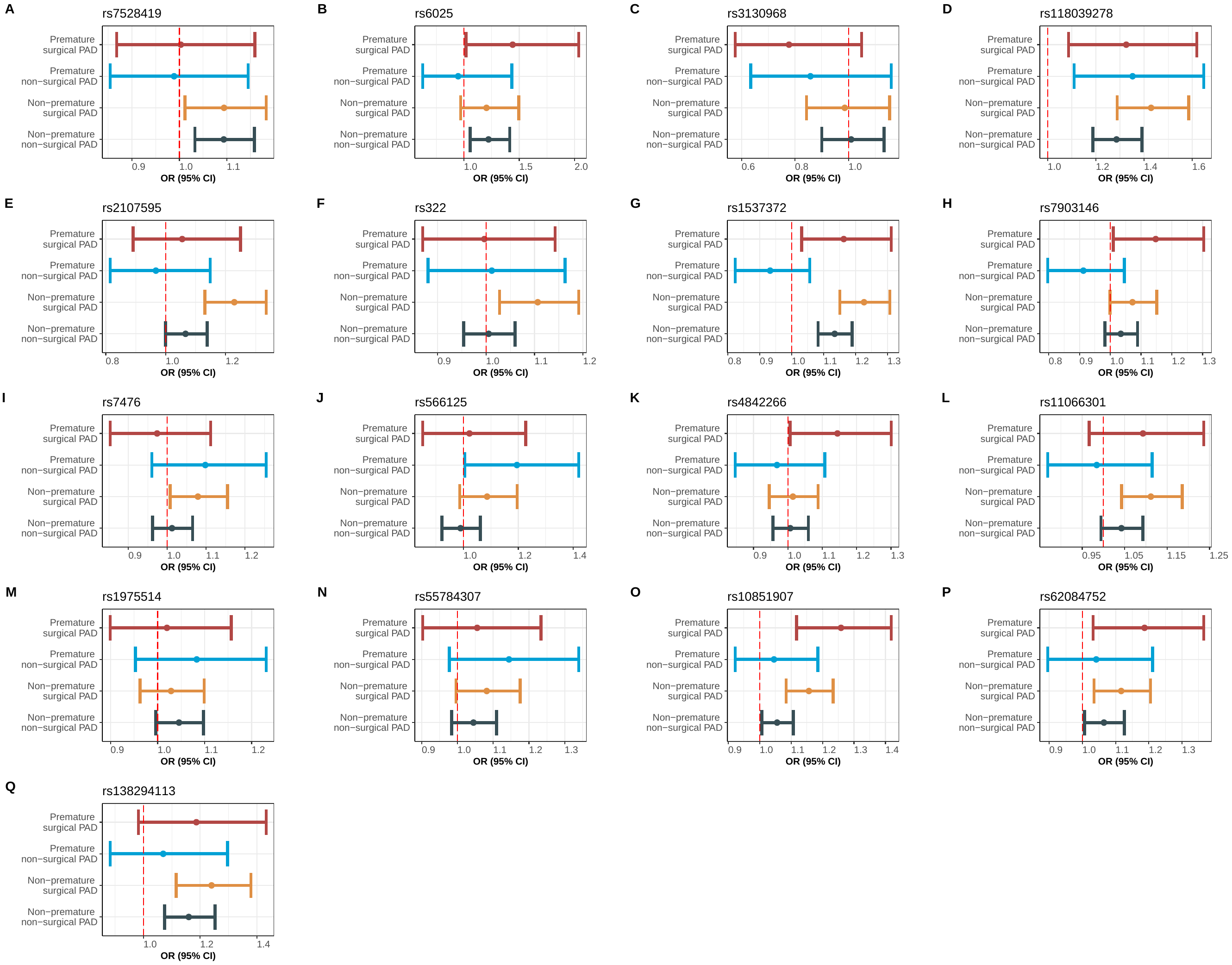

Results of 17 of 19 variants for the association analysis comparing four PAD severity subgroups with the non-PAD controls. Eight SNPs showed a higher OR in surgical PAD than in non-surgical PAD (E, G, H, K, L, O, P, Q).

**Figure S4 Distribution of PRS among PAD cases and non-PAD controls.**

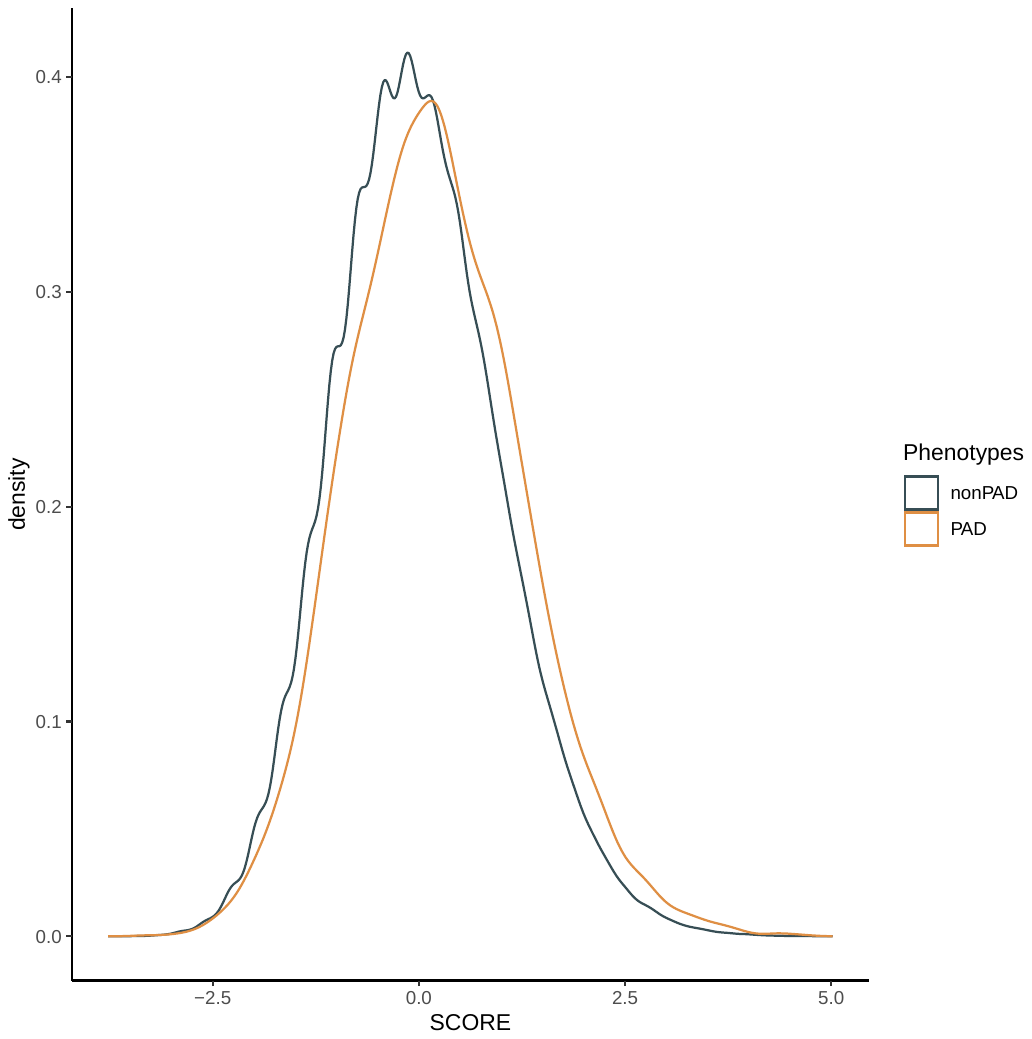

The standardized 19-variant PRS was higher in PAD cases than in non-PAD controls. The mean PRSs (standard deviations) were 0.209 (1.045) for PAD cases and -0.004 (0.999) for non-PAD controls.

**Figure S5 Receiver operating characteristic (ROC) curves for PRS predicting PAD and four severity subgroups.**

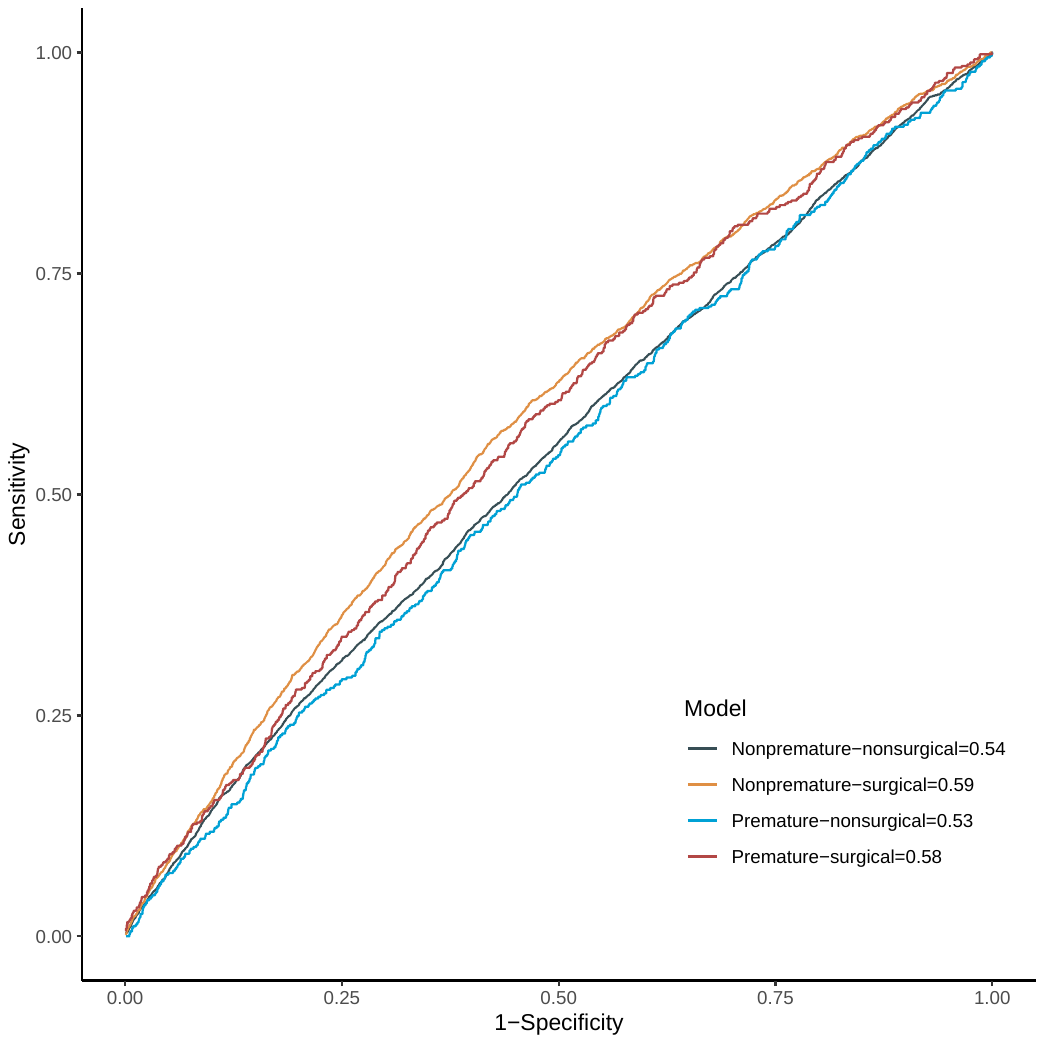

We used five-fold cross validation to develop and evaluate the performance of models of PRS predicting four severity subgroups compared with non-PAD controls. The Area Under Curves (AUCs) were computed, and the highest AUC was observed for Nonpremature-surgical PAD (0.59 95% CI: 0.56, 0.62).
